# Risky Sexual Behavior and its Determinants among Undergraduate Hostellers of Kathmandu Metropolitan City

**DOI:** 10.1101/2023.11.07.23298242

**Authors:** Ram Kumar Chaudhary, Anisha Chalise, Saloni Pandey, Shishir Paudel

## Abstract

**Background:** Risky sexual behavior (RSB) includes practices of unprotected sex, sex with multiple partners, and/or sex under substance abuse, increasing vulnerability to reproductive health problems. This study explored the risky sexual behavior and its associated factors among Nepalese undergraduates.

**Methods:** A cross-sectional study was executed among 361 undergraduates residing in hostels of Kathmandu Metropolitan. The data was collected through a self-administered questionnaire. Pearson’s chi-square test and multivariable logistic regression analysis was performed to determine factors associated with risky sexual behavior at 5% level of significance.

**Result:** The prevalence of risky sexual behavior among undergraduates was found to be 64.3% (95% CI: 59.8-69.8%). Risky sexual behavior was found to be associated with higher age (aOR: 3.938; 95% CI: 1.707-8.673), male gender (aOR: 3.233; 95% CI: 1.623-6.439), being in past/current relationship (aOR: 3.914, 95% CI: 2.099-7.012), lower education of mother (aOR: 3.655; 95% CI: 1.189-9.237), and peer pressure to have a sexual relationship (aOR: 2.356; 95% CI: 1.260-4.349). Notably, bivariate analysis illustrated problematic pornographic consumption to have a statistical relation with risky sexual behavior. However, this association weakened and became non-significant after accounting for other factors in the adjusted model (aOR:1.213, 95% CI: 0.331-4.442).

**Conclusion:** The study highlights a significant prevalence of risky sexual behaviour among undergraduate students, linked to the factors such as age, gender, relationship status, parental education, and peer pressure. These findings suggest the need for comprehensive sex education programs that equip students with the knowledge and skills to navigate healthy relationships, make safe choices, and embrace responsible sexual practices.

**Key Messages:** *What is already known on this topic:*

- Nepalese adolescents and youth have scant knowledge about sexual and reproductive health.
- Despite Nepal’s commitments to adolescent health, a gap persisted in the utilization of sexual and reproductive health services among youths and very little is known about their sexual practice.

*What this study adds:*

- Issues such as early sexual engagement, unprotected sex, multiple partners, and engagement in commercial sex work are prominent concerns among Nepalese youths.
- Despite existing laws and programs, risky sexual behaviors persist, emphasizing the urgency for effective interventions tailored to Nepalese adolescents.

*How this study might affect research, practice or policy:*

- The findings underscore urgent necessity for targeted interventions such as comprehensive sex education to address the concerning prevalence of RSB among Nepalese undergraduates.

## INTRODUCTION

Risky sexual behavior (RSB) is an act of getting engaged in unsafe sexual practices such as unprotected sexual intercourse, sex with multiple partners, and sex under substance abuse, making an individual vulnerable to reproductive health problems. Globally, an estimated one-million people encounter sexually transmitted infections, predominantly due to RSB and some through mother-to-child transmission.^1^ In the year 2022, globally, 39 million people were living with Human Immunodeficiency Virus (HIV) while 630 thousand deaths were accounted for AIDS-related illnesses.^2^ Since the start of the HIV epidemic, AIDS-related illnesses have claimed 40.4 million lives, with unsafe sexual practices recognized as a significant contributor for HIV & AIDS. ^1, 2^

Nepal is a signatory to the International Convention on Population and Development (ICPD), and has demonstrated its commitment to adolescent health by incorporating Adolescent Sexual and Reproductive Health (ASRH) into the National Reproductive Health Strategy since 1998.^3^ Despite the commitment, the utilization of sexual and reproductive health services remain unsatisfactory among adolescents and youth, mostly due to lack of adequate information about reproductive health.^4, 5^ A past study executed among adolescents attending a tertiary-level hospital revealed that 58.0% of the adolescents demonstrated only a moderate level of knowledge about sexual and reproductive health (SRH).^6^ Moreover, past studies consistently underscore the inadequate knowledge among Nepalese adolescents and youth regarding SRH practices.^7-9^ Scant knowledge of SRH during adolescence can make them vulnerable to unsafe sex and develop RSB in adulthood.

The Nepal Demographic and Health Survey of 2022 reported that only 16% of young women and 27% of young men possess thorough knowledge of HIV prevention methods. ^10^ Similarly, a study conducted in rural Nepal revealed that nearly two-third (73.2%) of school-going adolescents aged 15-24 years were engaged in various forms of RSB. ^11^ As undergraduates, individuals typically aged between 18 and 29 are at a crucial phase of life, making significant decisions and transitioning from adolescence to adulthood. This period is particularly vulnerable to the initiation of different lifestyle habits.^12^ Hostel residents, a distinctive group of students, living away from their home for educational purpose, can experience reduced parental guidance, making them more susceptible to adopt risky behaviors including RSB. Given the limited knowledge on the prevalence and nature of RSB among Nepalese youths, this study aims to explore the magnitude of this issue and its associated factors among Nepalese undergraduates.

## METHOD

### Study design

This was a cross-sectional study conducted among undergraduate students residing in the hostels of Kathmandu Metropolitan, Nepal, between July and October 2023.

### Participants

Undergraduate students residing in the sampled hostels for at least 6 months before data collection were eligible participants for the study. No exclusions were made based on age, gender, ethnicity, academic discipline, or other characteristics.

### Sample size determination and sampling technique

The sample size was determined using Cochran’s formula (n=z^2^pq/d^2^), where p referred to past prevalence of RSB (73%) observed among school-going students of rural Nepal aged 15 to 24 years^11^, q=1−p and d is the allowable error (5%). Considering 95% Confidence Interval (CI), the sample size was initially estimated at 301 which was optimized to 373 after adjusting for finite population of 6060 hostellers, and a 30% non-response rate, considering the sensitiveness of the research problem.

Initially, a total of 272 hostels within Kathmandu Metropolitan City registered under Nepal Hostel Association (NeHA) were contacted for the research permission, of which 196 hostels agreed for the support. From these, 20 hostels were randomly selected using STATS version 2.0 and the undergraduate students accommodating in these hostels were chosen randomly using lottery method.

### Data collection

The data were collected using self-administered questionnaire, distributed and collected from participants in a closed envelop in order to protect their anonymity. All the students were oriented about the questions presented in the questionnaire for clarification and written informed consent was acquired from them before handing them the enclosed questionnaire. Male participants were approached by male researcher and female participants were approached by female researcher. The questionnaire consisted of four sections. First section consisted of questions related to participants’ socio-demographic characteristics, followed by second section focusing on their lifestyles related attributes. Third second consisted of questions regarding their sexual behavior followed by Problematic Pornography Consumption Scale (PPCS),^13^ to assess their level of pornography exposure.

### Outcome Variable

The outcome variable for this study was risky sexual behavior which was assessed using a pretested tool prepared by the research team after an extensive literature review and in consultation with SRH experts. The tool, consisting of nine questions, explored risky sexual behaviors such as engaging in sex before the age of consent, having vaginal sex with intimate partner without using contraceptives even with no intention of pregnancy, engaging in vaginal sex with strangers without using condom, having oral sex with stranger or intimate partner even while sensing a risk of infection, not checking expiry date and/or damage of condoms, having multiple sex partners, having sex under the influence of alcohol, and having sex with commercial sex worker in past one year. A positive response to any of these questions was considered indicative of risky sexual behavior. The tool used for assessing risky sexual behavior is provided in Supplementary Document 1**(supp 1)**.

### Data processing, management and analysis

The collected data were carefully reviewed for accuracy and completeness. Data were entered in EpiData software V.3.1. A total of 10% of the randomly selected data were manually rechecked for accuracy in data entry and all data were exported to Statistical Package for the Social Sciences V.20 for statistical analysis. The data were summarized in terms of frequency, percentage, mean and standard deviation. The chi-square test and unadjusted odds ratio were calculated at 5% level of significance to identify the factors associated with risky sexual behavior. The factors found to have statistical significance (p<0.05) in chi-square test were subjected to the final model of multivariable logistic regressions for Adjusted odds ratio (aOR). Before performing multivariable analysis, the multi-collinearity among selected independent variables was tested using Variance Inflation Factor (VIF), where a VIF greater than five was taken as an indication of multi-collinearity between the independent variables.

### Patient and public involvement

None.

## RESULT

A total of 373 undergraduate hostellers were approached for this study, of which 361 provided their complete response to all questions. Thus, a response rate of 96.78% for all questions was acquired, and 361 samples were analyzed. The overall prevalence of risky sexual behavior was 64.3% (95% CI: 59.8-69.8). Out of 361 students, nearly one-third (37.7%) of them reported that they were sexually active in past one year prior to data collection. Among them, more than a quarter (28.7%) reported engaging in vaginal sex without contraceptives, even with no intent of pregnancy. Similarly, nearly one-third (33.1%) of the participants reported practicing oral sex with strangers without protection. Nearly a quarter (28%) reported having multiple sex partners, while nearly one-third (36%) reported engaging with commercial sex workers in past one year. (Table 1)

**Table 1:** Sexual Behavioral Practices among Undergraduate Students.

| **Characteristics** | **n** | **% (95% CI)** |
| --- | --- | --- |
| **Sexual Engagement** |  |  |
| No | 225 | 62.3 (57.6-67.3) |
| Yes | 136 | 37.7 (32.7-42.4) |
| **Age at first sexual intercourse (n=136)** |  |  |
| ≥18 years | 113 | 83.1 (76.5-89.7) |
| ≤17 years | 23 | 16.9 (10.3-23.5) |
| **Vaginal sex without contraceptives (n=136)** |  |  |
| Yes | 97 | 71.3 (62.8-78.3) |
| No | 39 | 28.7 (21.7-37.2) |
| **Oral sex without protection (n=136)** |  |  |
| Yes | 45 | 33.1 (25.3-40.8) |
| No | 91 | 66.9 (59.2-74.7) |
| **Vaginal sex with stranger without condom (n=136)** |  |  |
| Never | 59 | 43.4 (36.0-53.7) |
| Sometimes | 66 | 48.5 (39.7-57.8) |
| Often | 11 | 8.1 (4.0-13.2) |
| **Check expiry dates and/or damage of condom (n=136)** |  |  |
| Never | 60 | 44.1 (35.6-52.9) |
| Sometimes | 27 | 19.9 (14.7-26.5) |
| Often | 49 | 36.0 (27.9-43.4) |
| **Number of sex partners (n=136)** |  |  |
| Single partner | 98 | 72.0 (63.9-78.7) |
| More than one partner | 38 | 28.0 (8.8-31.3) |
| **Engaged in sex under influence of alcohol (n=136)** |  |  |
| Never | 79 | 58.1 (50.3-65.8) |
| Sometimes | 49 | 36.0 (27.9-45.3) |
| Often | 8 | 5.9 (2.2-9.6) |
| **Sex with Commercial sex workers (n=136)** |  |  |
| Never | 87 | 64.0 (55.5-72.5) |
| Once | 21 | 15.4 (8.4-21.3) |
| Twice | 17 | 12.5 (8.1-18.8) |
| Three or more times | 11 | 8.1 (3.7-12.5) |
| **Overall Sexual Practice** |  |  |
| Risky | 232 | 64.3 (59.8-69.8) |
| Non-Risky | 129 | 35.7 (30.2-40.2) |

The participant’s age ranged from 18 to 32 years, with a mean age of 21.7±2.72 years. Among the 361 participants, the largest group of participants (29.9%) were enrolled in the management stream, followed by health sciences (28.8%), education/humanities (21.1%), engineering (11.6%) and basic sciences (8.6%). Slightly more than half (57.1%) of the participants were male. Nearly half (55.7%) reported being in relationship currently or within one year preceding data collection. In bivariate analysis, socio-demographic factors such as participants’ age, gender, relationship status and educational level of both parents were found to be associated with RSB. (Table 2)

**Table 2:** Association of Risky sexual behavior with socio-demographic Characteristics.

| Characteristics | n (%) | Risky Sexual Behaviour | | $\chi^2$ | p-value |
| --- | --- | --- | --- | --- | --- |
|  |  | Presence<br>n (%) | Absence<br>n (%) |  |  |
| **Age** |  |  |  |  |  |
| <20 years | 133 (36.8) | 34 (25.6) | 99 (74.4) | 18.489 | <0.001* |
| 20 to 25 years | 178 (49.3) | 65 (36.5) | 113 (63.5) |  |  |
| $\geq 25$ years | 50 (13.9) | 30 (60.0) | 20 (40.0) | | |
| **Gender** |  |  |  |  |  |
| Male | 206 (57.1) | 104 (50.5) | 102 (49.5) | 45.462 | <0.001* |
| Female | 155 (42.9) | 25 (16.1) | 130 (83.9) |  |  |
| **Family type** |  |  |  |  |  |
| Nuclear | 213 (59.0) | 69 (32.4) | 144 (67.6) | 2.523 | 0.112 |
| Joint/extended | 148 (41.0) | 60 (40.5) | 88 (59.5) |  |  |
| **Relationship status** |  |  |  |  |  |
| Never been in relationship | 160 (44.3) | 30 (18.8) | 130 (81.3) | 36.095 | <0.001* |
| Been in relationship (Past/Current) | 201 (55.7) | 99 (49.3) | 102 (50.7) |  |  |
| **Education status of father** |  |  |  |  |  |
| Illiterate | 17 (4.7) | 11 (64.7) | 6 (35.3) | 9.659 | 0.022* |
| Literate with informal education | 20 (5.5) | 8 (40.0) | 12 (60.0) |  |  |
| Up to secondary level (1-10 grade) | 134 (37.1) | 53 (39.6) | 81 (60.4) |  |  |
| Higher education | 190 (52.7) | 57 (30.0) | 133 (70.0) |  |  |
| **Education Status of mother** |  |  |  |  |  |
| Illiterate | 38 (10.5) | 21 (55.3) | 17 (44.7) | 8.652 | 0.034* |
| Literate with informal education | 52 (14.4) | 21 (40.4) | 31 (59.6) |  |  |
| Up to secondary level (1-10 grade) | 156 (43.2) | 53 (34.0) | 103 (66.0) |  |  |
| Higher education | 115 (31.9) | 34 (29.6) | 81 (70.4) |  |  |
| **Current academic status** |  |  |  |  |  |
| Health science | 104 (28.8) | 30 (28.8) | 74 (71.2) | 6.435 | 0.169 |
| BSc science | 31 (8.6) | 16 (51.6) | 15 (48.4) |  |  |
| Management | 108 (29.9) | 40 (37.0) | 68 (63.0) |  |  |
| Engineering | 42 (11.6) | 13 (31.0) | 29 (69.0) |  |  |
| Education/Humanity | 76 (21.1) | 30 (39.5) | 46 (60.5) |  |  |
| **Academic year** |  |  |  |  |  |
| First year | 128 (35.5) | 38 (29.7) | 90 (70.3) | 6.023 | 0.110 |
| Second year | 93 (25.8) | 38 (40.9) | 55 (59.1) |  |  |
| Third year | 65 (18.0) | 20 (30.8) | 45 (69.2) |  |  |
| Fourth year | 75 (20.8) | 33 (44.0) | 42 (56.0) |  |  |

\* Significance at $p < 0.05$

Out of 361 participants, nearly one-fifth (19.4%) reported belonging to a rural community. Approximately one-third (29.6%) experienced peer pressure to engage in a sexual relationship. Assessment using the Problematic Pornography Consumption Scale revealed that 12.7% exhibited behaviors suggesting a risk of problematic pornography consumption, scoring between 50-74 in PPCS, while 4.4% met the criteria for problematic consumption with a PPCS score>75. Bivariate analysis indicated a statistically significant relationship of peer pressure for sexual relationships, smoking, and pornography consumption with RSB. (Table 3)

**Table 3:** Association of Risky Sexual Behavior and lifestyle and psychological factors.

| Variables | n (%) | Risky Sexual Behaviour | | $\chi^2$ | p-value |
| --- | --- | --- | --- | --- | --- |
|  |  | Presence<br>n (%) | Absence<br>n (%) |  |  |
| **Home town** |  |  |  |  |  |
| Urban | 291 (80.6) | 104 (35.7) | 187 (64.3) | 0.000 | 0.997 |
| Rural | 70 (19.4) | 25 (35.7) | 45 (64.3) |  |  |
| **Peer pressure for sexual relationship** |  |  |  |  |  |
| Absence | 254 (70.4) | 61 (24.0) | 193 (76.0) | 51.242 | <0.001* |
| Presence | 107 (29.6) | 68 (63.6) | 39 (36.4) |  |  |
| **Smoking status** |  |  |  |  |  |
| No smoking | 299 (82.8) | 88 (29.4) | 211 (70.6) | 30.114 | <0.001* |
| Smoking | 62 (17.2) | 41 (66.1) | 21 (33.9) |  |  |
| **Victim of sexual abuse** |  |  |  |  |  |
| No | 330 (91.4) | 123 (37.3) | 207 (62.7) | 3.962 | 0.050 |
| Yes | 31 (8.6) | 6 (19.4) | 25 (80.6) |  |  |
| **Status of pornography consumption** |  |  |  |  |  |
| Non-problematic consumption | 299 (82.8) | 97 (32.4) | 202 (67.6) | 8.968 | 0.011* |
| At risk of problematic consumption | 46 (12.7) | 22 (47.8) | 24 (52.2) |  |  |
| Problematic consumption | 16 (4.4) | 10 (62.5) | 6 (37.5) |  |  |

\* Significance at $p < 0.05$

**Table 4:**
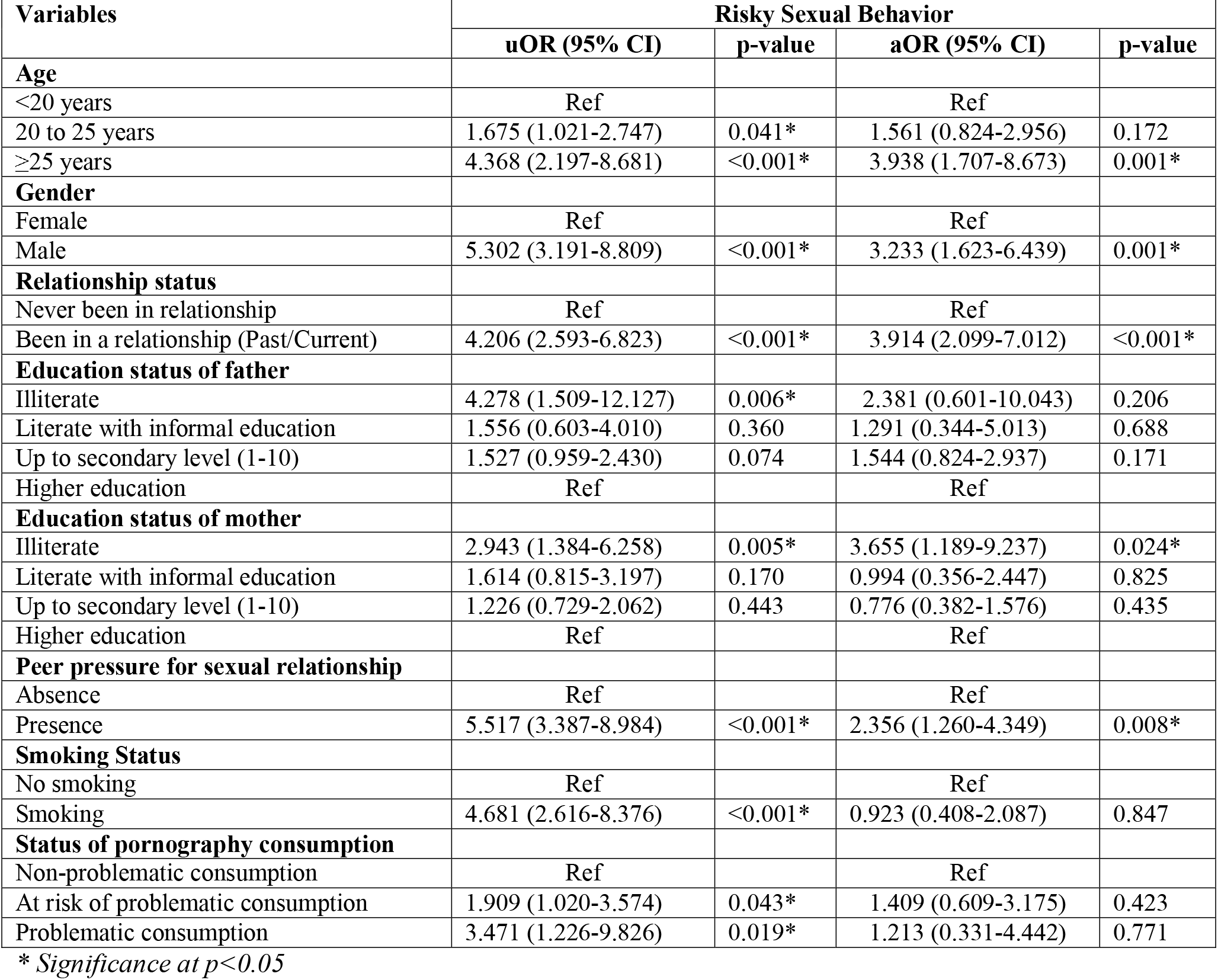
Factors associated with Practice towards Sexual Behavior.

For multivariate analysis, the VIF test was performed among the independent variables which were found to have statistically significant relationship with RSB in bivariate analysis. The highest reported VIF was 1.749, confirming no issue of multi-collinearity. It was observed that compared to undergraduates under 20 years, those above 24 were thrice more at odds (aOR: 3.938; 95% CI: 1.707-8.673) of practicing RSB. Similarly, males were observed to have three times higher odds (aOR:3.233; 95% CI: 1.623-6.439) of RSB as compared to females. Individuals in past or current relationships had a three-fold increase in odds (aOR: 3.914, 95% CI: 2.099-7.012) of practicing RSB as compared to those never in a relationship. Participants whose mothers had low educational attainment had higher odds of RSB (aOR: 3.655; 95% CI: 1.189-9.237). It was observed that experiencing peer pressure to have a sexual relationship could double the odds of RSB (aOR: 2.356; 95% CI: 1.260-4.349). Notably, bivariate analysis illustrated that participants with problematic pornographic consumption were three times more at odds of practicing RSB. However, this association weakened and became non-significant after accounting for other factors in the adjusted model (aOR: 1.213, 95% CI: 0.331-4.442).

## DISCUSSION

This study assessed the practice of risky sexual behavior along with its associated factors among undergraduate hostel students of Kathmandu Metropolitan city. It was noted that nearly three out of five (64.3%) participants were engaged in risky sexual behavior. This proportion of risky sexual behavior is slightly lower than the prevalence shown by a past study from rural Nepal where 73.2% of the sexually active participants were found practicing RSB.^11^ However, another study focusing on RSB among young men in Nepal reported 20% of sexually active single men and 9% married men to be engaged in RSB.^14^ The practice of RSB has been reported among undergraduates throughout the world from different countries such as Sri Lanka, Ethiopia, and Rwanda where the risky sexual behavior among undergraduates was observed to be between 12.1% and 64.0%. ^15-17^ Similarly, a meta-analysis based on eighteen studies from Ethiopia estimated the pooled prevalence of RSB among college and university students to be 41.62%.^18^ The variation in prevalence of RSB might be attributable to the difference in sample size and study setting as well as the variation in case definition of RSB in these studies. Despite the variations, these observed rates of RSB indicates risky sexual practice among youth is a concerning public health issue.

Among the various RSB, prominent issues include engaging in sexual activity before reaching a legal age, practicing unprotected vaginal and oral sex, having multiple sexual partners, neglecting condom quality checks, and sexual engagement with commercial sex workers. In line with this observation, a study based on Pokhara Metropolitan observed that 85.09% of the students had their sexual experience between the age of 15-19 years, while 10.56% were sexually active before the age of 15 and 13.33% adolescents had unprotected sex.^19^ Similarly, the initiation of sexual engagement before the age of 18 and unprotected sex has been reported by multiple studies from Nepal.^20, 21^ Likewise, a study from rural Nepal also reported that 28.2% of the sexually active youth were engaged with multiple sexual partners.^11^ Engagement with multiple sexual partners has been reported by other studies as well.^19, 20^ Similarly, it was also observed that nearly two-third (36%) of the undergraduates had sex with commercial sex worker, despite the fact that commercial sex work has been considered as an act of trafficking in Nepal and is punishable under Human Trafficking and Transportation (Control) Act 2008, penalizing those involved directly or indirectly in prostitution, which is also endorsed by National Penal Code, 2074.^22^ The sexual engagement of students with commercial sex workers has also been reported by numerous studies from Nepal where it is expected to range between 8.5% and 48.9%.^11, 19-21, 23^ Despite presence of laws and programs aimed to prevent minors’ engagement in sexual activities to protect them from sexual abuse, as well as regulations controlling commercial sex work and various initiatives focusing on adolescent SRH, these risky behaviors persist. This underscores the urgent necessity for more effective and comprehensive sexual education interventions among Nepalese adolescents and youth. It is crucial to enhance their understanding of SRH and reduce the risks of sexually transmitted infections within this vulnerable group.

It was observed that the odds of RSB increased with the increase in age of the participants. This is in line with a study from Ethiopia where students above the age of 24 years were found to be twice more likely to engage in RSB as compared to younger students.^16^ This finding aligns with several cross-sectional studies that have explored the complex relationship between age and sexual behavior and observed that age may play a crucial role in influencing individuals’ likelihood of engaging in RSB.^18, 20, 24, 25^ A plausible explanation for this association could be that the students of higher age may be more inclined to be involved in relationships, exhibit greater confidence in making independent decisions, and enjoy improved financial options compared to their younger counterparts.

Males were thrice more likely to be engaged in RSB than females. This is in line with other studies from Nepal, where males were reported to express more sexual engagements and higher risk of sexual behavior.^11, 20^ Globally, studies have reported higher proportion of RSB among males as compared to females.^17, 18, 25^ Studies noted that males tend to report higher sexual exposure than females,^15^ which might be the case in our study as well. The difference of RSB across gender could be attributed to many factors such as social desirability biases and sexual double standards such as women are expected to be virgin till marriage, and male’s masculinity is associated with sex. This might be the reason that male are more likely to engage with commercial sex workers, have multiple partners and express their sexual escapades in comparison to females.^20^ Similarly, students with mothers having lower education were found to have three fold increase in odds of RSB. Studies have stated that family environment and parental education could play a significant role in influencing sexual behavior in later life.^20, 25^ These observations suggest that addressing gender-specific social norms and enhancing parental education may prove effective in mitigating RSB among youth.

Participants who were in relationships were thrice more likely to engage in RSB. Past studies have shared that the first sexual partner in most cases are often a boyfriend or girlfriend.^15, 23^ Additionally, it has also been observed that students who are in relationship are more likely to get engaged in premarital sexual relationship,^20^ thereby increasing their risk of RSB. A study based on African college students partially supported the hypothesis that students in long-term relationships were more likely to engage in RSB.^26^ Likewise, it was also observed that students experiencing peer pressure to have sexual relationship were twice more likely to experience RSB. This is in line with a study based on Kathmandu, which suggests that peers play an important role in influencing the views, attitudes, and sexual behavior of individuals.^23^ Peer pressure has been observed to be one of the important factor for RSB among students in many counties.^17^ Thus, enhancing comprehensive sex education programs is crucial to promoting safer sexual practices among this vulnerable group and addressing the influence of peers and peer pressure on risky sexual behavior.

Bivariate analysis revealed that participants with problematic pornographic consumption were three times more likely to engage in RSB, which is in line with past observations,^18^ where pornographic exposure has been thought to increase the motivation of sexual desire. It has been highlighted that exposure to sexually explicit media contents including pornography in early adolescence can be a predictor for RSB such as early sexual engagement, unsafe sex, and multiple sexual partners.^27^ However, in this study, this association weakened and became non-significant after accounting for other factors in the adjusted model. This is in line with another observation which suggest that early exposure to sexually explicit material could be additive risk factor for sexual risk taking but pornography might not completely contribute to sexual risk taking among young adults.^24^ This suggests that while problematic pornography consumption might be linked to risky sexual behavior, other factors play a more crucial role once accounted for.

Despite being one of the few studies exploring the RSB among undergraduates of Nepal along with its risk factors, it is crucial to acknowledge its inherent limitations, and the findings should be interpreted based on these limitations. Although we tried to make the data collection session as private and confidential as possible, due to the sensitive nature of the research topic and self-reported approach, there may be slight possibility of social desirability bias as some students could have deliberately hidden some information in relation to unacceptable behavior. Despite Kathmandu being the national capital housing diverse Nepalese citizens from all demographic backgrounds and our effort to include students from various academic streams for improved generalizability, it is important to note that Nepal, being a small yet culturally rich and diverse nation, the study area may not fully encompass all aspects of the nation’s cultural, racial, and ethnic diversity. Thus, a larger community-based survey covering these factors might provide additional insights.

## Conclusion

The study underscores a significant prevalence of risky sexual behaviour among undergraduate hostellers of Kathmandu, revealing concerning trends such as early sexual engagement, unprotected sex, multiple partners, and engagement in commercial sex work. The findings highlight the urgent need of comprehensive sexual education interventions tailored to the specific needs of Nepalese adolescents and youth, focusing on social norms, gender-specific education, and parental involvement, emphasizing the importance for reducing risks associated with sexually transmitted infections within this vulnerable group.

## Data availability statement

The dataset generated and analyzed during the current study is available from the corresponding author upon reasonable request.

## Ethics statements

### Patient consent for publication

Not applicable.

### Ethics approval

This study involves human participants and ethical approval for this study was obtained from the institutional review committee of CiST College (Ref no. 47/080/081). Formal approval was acquired from the Hostel Association and selected hostels. Written inform consent was obtained from each participant prior to data collection. Questionnaire was distributed to and collected back from participants in a closed envelop, in order to maintain confidentiality of participant’s information. To ensure privacy, the participants were requested to fill the questionnaire in an empty room of their hostel.

## Acknowledgements

The authors would like to thank all the undergraduate students who participated in this study and provided their valuable time and information. Without their support, this study would not have been possible. The authors would also like to extend their gratitude towards Nepal Hostel Association and selected hostels for their approval and constant support during data collection.

## Contributors

RKC, AC and SP conceptualized the study. RKC and SP developed questionnaire. RKC and SaP were engaged in data collection. Statistical analysis was planned and carried out by RKC and SP, with contributions and review from AC, SaP. SP and AC supervised the overall work and wrote first draft. All authors contributed to data interpretation, reviewed successive drafts, and approved the final version of the manuscript.

